# Multi-task deep learning-based survival analysis on the prognosis of late AMD using the longitudinal data in AREDS

**DOI:** 10.1101/2021.08.26.21262548

**Authors:** Gregory Ghahramani, Matthew Brendel, Mingquan Lin, Qingyu Chen, Tiarnan Keenan, Kun Chen, Emily Chew, Zhiyong Lu, Yifan Peng, Fei Wang

## Abstract

Age-related macular degeneration (AMD) is the leading cause of vision loss. Some patients experience vision loss over a delayed timeframe, others at a rapid pace. Physicians analyze time-of-visit fundus photographs to predict patient risk of developing late-AMD, the most severe form of AMD. Our study hypothesizes that 1) incorporating historical data improves predictive strength of developing late-AMD and 2) state-of-the-art deep-learning techniques extract more predictive image features than clinicians do. We incorporate longitudinal data from the Age-Related Eye Disease Studies and deep-learning extracted image features in survival settings to predict development of late-AMD. To extract image features, we used multi-task learning frameworks to train convolutional neural networks. Our findings show 1) incorporating longitudinal data improves prediction of late-AMD for clinical standard features, but only the current visit is informative when using complex features and 2) “deep-features” are more informative than clinician derived features. We make codes publicly available at https://github.com/bionlplab/AMD_prognosis_amia2021.

## 1 Introduction

Age-related macular degeneration (AMD) is the leading cause of vision loss, and is projected to affect approximately 288 million people by 2040^1–3^. In the United States alone, the annual healthcare cost of treating this disease is $4.6 billion, creating an extreme burden on patients and the healthcare system^4^. AMD is characterized by the destruction of a retinal pigment epithelial (RPE) cells, which directly interact with photoreceptors to allow for proper function of the eye^5^. In AMD, drusens, or lipid deposits, form near the RPE cells, which can eventually lead to tissue atrophy in the eye. In addition, RPE cells normally contain melanosomes, which create a certain pigmentation in the eye.

The onset of the disease can be heterogeneous between individuals. The majority of patients have a form of the disease known as dry AMD, which has fairly slow progression, whereas some patients (10-15% of early-stage AMD) will develop choroidal neovascularization (CNV) which leads to the rapid loss of vision and faster onset of late-stage AMD^5^. Therefore, to improve treatment plans for patients, it can be useful to understand the risk of developing CNV and in particular, the risk of developing late-stage AMD.

The current method to assess AMD severity requires the use of color fundus photographs (CFP), which are generated by a low-power microscope that captures general eye health and examines structures within the eye^6^. These photographs are then sent to grading centers, where experts analyze specific characteristics, including presence, type, and extent of drusens, presence/extent of retinal depigmentation, serous sensory retinal detachments, subretinal hemorrhages, subretinal fibrosis, and geographic atrophy, which are used for characterizing AMD severity. A simplified AMD severity score and risk classification has been developed by the Age-Related Eye Disease Study (AREDS) Research Group^7^. Based on characteristics from CFPs at the current time of visit, patients are binned into 5 categories (0-4), which estimate the likelihood the patient will progress to late-stage AMD. This five-step simplified severity scale is the current clinical standard in assessing a patient’s risk of developing late-AMD. This risk is calculated using the size of drusens, presence of pigmentation abnormalities, age, and smoking status at the current time of visit.

Over the past decade, the use of deep learning has grown exponentially. Convolutional neural networks (CNNs) have been used to identify patterns within images to classify medical imaging data. Various models have been developed to characterize CFPs, based on several characteristics. Single task models have been used to classify characteristics such as geographic atrophy and drusen presence^8,9^. In addition, multi-task models have been developed to characterize these eye characteristics simultaneously^10^. Subsequently, researchers have used these image features derived from a CNN model in a survival setting to predict patients who are at risk of developing late-stage AMD^11^. However, it is well known that the rate of progression for patients within the early-stage AMD category is heterogeneous. In this work we hope to utilize the time-varying information for these patients to improve upon risk-prediction for AMD patients.

We combine elements of multiple past works to improve upon AMD patient stratification, while introducing a novel time-varying component to improve model performance. A multi-task learning model was used to predict both drusen size and presence of pigmentation abnormalities in the right and left eyes of patients. Drusen size and presence of pigmentation abnormalities are the criteria used for the simplified AREDS severity scale. Image features are extracted from the multi-task learning model. Multi-task learning was incorporated to extract more generalizable image features than the clinician derived features, hoping to improve our ability to predict risk of developing late-AMD. Either the image features or clinical features are passed through either a multilayer-perceptron (MLP) or long short-term memory (LSTM) network, to predict patient risk of developing late-stage AMD. A survival loss function is utilized to train the risk prediction model to account for patients that end the study without developing late-stage AMD. We compare model performances of these image derived features with those of expert-derived features to compare our results to a baseline model. The end goal of this work is to (a) reduce the burden on grading centers by reducing the time needed to analyze simple cases and assisting in edge-case classification and (b) use the features derived from the images to improve upon the stratification of patients with early-stage AMD based on the risk of progressing to late-AMD.

## 2 Materials and Methods

### 2.1 Dataset

In this study, we use the AREDS cohort sponsored by the National Eye Institute (National Institutes of Health). It was a 12-year multi-center prospective cohort study of the clinical course, prognosis, and risk factors of AMD, as well as a phase III randomized clinical trial to assess the effects of nutritional supplements on AMD progression^12^. In short, 4,757 participants aged 55 to 80 years were recruited between 1992 and 1998 at 11 retinal specialty clinics in the United States. The inclusion criteria were wide, from no AMD in either eye to late AMD in one eye. The AREDS dataset is publicly accessible to researchers by request at dbGAP^c^. In the AREDS cohort, at baseline and at annual visits, comprehensive eye examinations were performed by certified study personnel using a standardized protocol, and CFP (field 2, i.e., 30° imaging field centered at the fovea) were captured by certified technicians using a standardized imaging protocol.

The longitudinal analysis of the AREDS cohort led to the development of the patient-based AREDS Simplified Severity Scale for AMD, based on the grading of CFP^7^. This simplified scale provides convenient risk factors for assessing the risk of progression to late AMD that can be determined by clinical examination or by less demanding photographic procedures than used in the AREDS. The scale combines risk factors from both eyes to generate an overall score for the individual, based on the presence of one or more large drusen (diameter > 125 mm) and/or AMD pigmentary abnormalities at the macula of each eye. The Simplified Severity Scale is clinically useful in that it allows ophthalmologists to predict an individual’s 5-year risk of developing late AMD. This 5-step scale (from score 0 to 4) estimates the 5-year risk of the development of late AMD in at least one eye as 0.4%, 3.1%, 11.8%, 25.9%, and 47.3%, respectively.

In our study, the event of interest was the development of late AMD. The ground truth labels (AREDS Simplified Severity Scale and phenotype features such as drusen status and macular pigmentary abnormalities) used for both training and testing were the grades previously assigned to each CFP by expert human graders at the University of Wisconsin Fundus Photograph Reading Center. The reading center workflow has been described previously^13^.

To train and test our models, we created a data subset that consisted of 3,747 patients from AREDS who had not reached late AMD through year 3. Of these patients, 2.7% reached late-AMD by year 5, 6.1% reached late-AMD by year 8, and 9.3% reached late AMD by the end of the study. This dataset consisted of the gradings from the certified study personnel, as well as inferred grades, such as presence of one or more large drusen and AMD pigmentary abnormalities at the macula of each eye.

### 2.2 Development of the algorithm

Figure 1 shows the overarching architecture used to A) develop the fine-tuned CNN drusen size and pigmentation abnormality classifiers trained on images from all patients and B) extract the “fine-tuned” and “pretrained” features on the images from years 0, 2, and 3 for the patients who had not reached late-AMD by year 3. Figure 1C demonstrates example model of how image features are used to predict risk of developing late-AMD at years 5, 8, and overall risk. We will describe each module in the subsequent sections.

**Figure 1:**
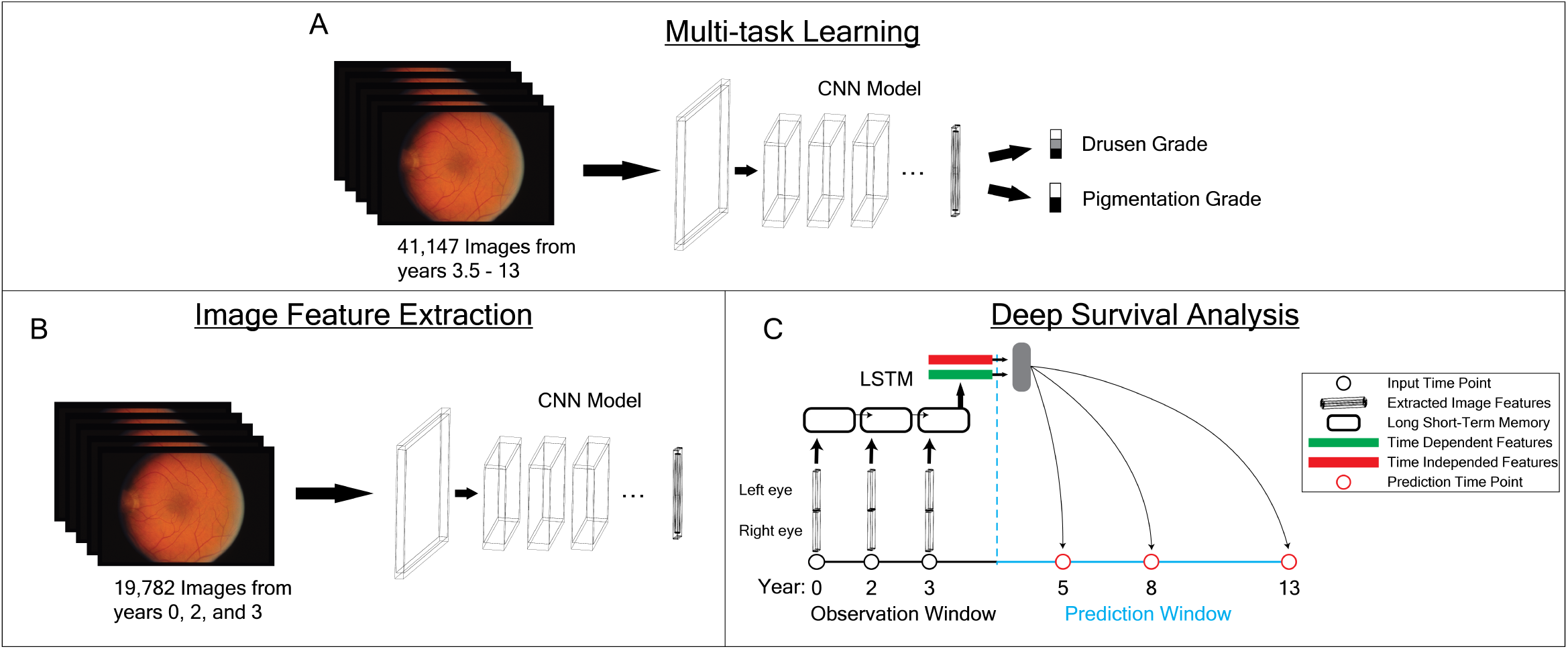
Model Architecture.

#### 2.2.1 Multi-task Learning

Multi-task learning is a field of machine learning where multiple tasks are learned in parallel while using a shared representation^10,14,^15. It exploits the similarities (shared image features) and differences (task-specific image features) between the features present on different tasks, thus reduces the losses of various tasks simultaneously.

In this study, we created a multi-task deep learning model that trains the classification of drusen size and presence of pigmentation abnormalities simultaneously. Drusen size and pigmentation abnormalities are the features used to calculate the 5-step simplified severity scale. Of the 60,929 CFPs used in this study, 41,147 images from visits after year 3 were randomly sampled in a 90-10 split to train and test. Data augmentation was performed to improve model generalizability. Training images were randomly horizontally flipped, cropped, blurred, rotated, sheared, morphed, and the contrast was randomly strengthened or weakened using the ImgAug module in Python^d^. These augmentations were set to plausible realistic ranges of CFPs. All images were resized to 256 × 256, then center cropped to 224 × 224. This aided in removing the unwanted areas as there is some blank space surrounding the eye image. Images were then normalized to a mean of 0 and standard deviation of 1 based on the mean and standard deviation of 10,000 randomly sampled images from the entire dataset.

Two different models were trained for this study to compare performance. ResNet152^16^ and EfficientNetB3^17^ were used to see how deep learning architecture affects model performance. Weights on both the ResNet152 and EfficentNet-B3 models were both initialized to the pretrained ImageNet weights^18–20 e^. The last-fully-connected layers of both models were replaced with two separate linear layers, one for predicting size of drusen (three-class) and one for predicting pigmentation abnormality (two-class) (Figure 1A).

To account for the class imbalance, weighted cross entropy loss was used on each classifier. Weightings were set to 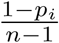, where p_*i*_ is the proportion of class *i* in the training set, and n is the number of classes. The model loss was set to the mean of the drusen classifier loss and the pigmentation classifier loss. The Adam algorithm was used as an optimizer with the learning rate set to 0.005. Up to 25 epochs, with a batch size of 16 images were run to train each model. Training ceased when the training loss dropped below 85% of the test loss. The weights from the epoch with the lowest test loss were saved and used to extract image features for deep survival analysis.

### 2.3 Image Feature Extraction

To extract image features, the fully-connected layers from the multi-task learning model were removed, and an n-dimensional vector was extracted from the last hidden layer for each image (2,048 for ResNet152 and 1,536 for EfficientNetB3). These features will be referred to as “fine-tuned” features. A comparison is also done with the ImageNet pretrained model to demonstrate improved feature extraction using the multi-task learning model. Those features will be referred to as “pretrained” features. Collectively, they are referred to as “deep” features.

### 2.4 Survival Analysis

Deep image features from CFPs at years 0, 2, and 3 and clinical data of 3,297 patients who had not reached late-AMD at year 3 were used to predict risk of developing late-AMD at years 5 and 8 and by the end of the study. Five-fold cross validation was used to evaluate the performance of all models. Of the 80% of the patients not in the test set, 80% were used to train the model and 20% were used as a validation set. One single batch was used during training to account for low numbers of uncensored data.

#### CoxPH Model

Image features from each eye and each visit were concatenated to an *n* × 2 × *m* vector, where *n* is the size of the CNN output (2048 for ResNet152 and 1536 for EfficientnetB3) and *m* is the number of years included. Using principal component analysis (PCA) from the scikit-learn library, a linear transformation to 10 dimensions was made on the training dataset^21^. On average, these PCA decompositions explained 22.4%, 85.4%, 81.6%, and 73.6% of the variance for the ResNet152 pretrained, ResNet152 fine-tuned, EfficientNetB3 pretrained, and EfficientNetB3 fine-tuned feature vectors across the cross-validation training sets, respectively. Fewer than 10 principal components further reduced the explained variance, while more than 10 led to linearity convergence issues with the CoxPH model. The CoxPH Fitter model from the lifelines module was then fit with a step size of 0.1 and no penalizer, using the Breslow method for handling ties^22f^.

#### MLP Model

For each patient, deep image features were extracted from images of the patient’s left eye and right eye at years 0, 2, and 3. Features were z-score normalized, fit to the training set, prior to training and evaluating the survival models. The models incorporated time-dependent information in two different ways, both utilizing the pytorch deep learning library^20^. For the MLP model, the left and right eye image features were concatenated for the three different time points, generating a (*n* × 2 × *m*) dimensional vector. The MLP model consisted of one hidden layer (32-dimension for 1 year and 96-dimension for 3 years) that then fed into a final linear layer with one node as the output for the survival loss calculation.

#### LSTM

For the LSTM model (Figure 1C), the left and right eye data for each time point were concatenated separately for each visit, then passed through a single linear layer to decrease the dimensionality of the concatenated feature vector by a factor of 8, creating a tensor of size (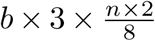), where *b* is the batch size and *n* in the size of the feature vector from the CNN. Each of these time points were then used as a separate input into the LSTM to model the time-varying changes in eye features. The hidden state from the final time point was fed into a survival loss similar to what was done for the MLP model. The hidden state size for the model was 128. In addition, dropout (*p* = 0.6) was used prior to the linear layer to reduce overfitting. All models used the pycox loss, which is an approximation of the negative partial log likelihood^23^. We adopted survival loss using Efron’s method to handle ties in the survival time using code from the Pysurvival package^24^.

### 2.5 Evaluation Metrics

#### Survival Analysis

To evaluate model performance, several different metrics were used. Concordance index was calculated using the pycox package^25^. In addition, patients were categorized into late-stage or not late-stage AMD at 2 years (year 5) and 5 years (year 8) after the year 3 time point. We then generated area under the receiver operating characteristic curve (ROC AUC) for predicting late-stage AMD at years 5 and 8 based on the risks generated from our model (represented as ROC AUC@5 and ROC AUC@8)^26^. In addition, using the R-based timeROC package with the rpy2 package, we evaluated the precision and recall of the models by generating AUC values indicating the extent of false positives and false negatives in the prediction^g^. To visualize classification performance of the CNN model and to examine how they correlate with clinical features, we used t-SNE from the scikit-learn package in Python^h^. Matplotlib was used for plotting all analyses^i^.

#### Multi-task Learning Analysis

We constructed contingency tables of the true and predicted values of drusen size and pigmentation abnormalities for both the fine-tuned ResNet152 and EfficientNetB3 models. Overall accuracy, sensitivity, specificity, and precision were calculated for each class. Drusen sensitivity, specificity, and precision were calculated in a one-vs-all method, where the identified value was considered positive and the other two values considered negative.

### 2.6 Analyzing Clinical Features

Two distinct datasets were used with both linear and non-linear models to predict the risk of patients developing late-AMD. The first set, labeled as clinical set A, contained age, smoking status, and for each eye, drusen size and presence of pigment abnormalities. These are the features used to calculate the 5-step simplified severity scale. Clinical set B contained age, smoking status, and for each eye: area of drusens within a central grid supplied to the grader, geographic atrophy (GA) within the central grid, subretinal GA atrophy, subretinal fibrosis, non-drusenoid pigment epithelial detachment, serous sensory retinal hemorrhaging, subretinal or subRPE hemorrhaging, RPE depigmentation, and increased pigmentation within the central grid. Drusen suze and presence of pigmentation abnormalities in dataset A are calculated by binning features in dataset B to offer a more immediate and interpretable interpretation of the wellbeing of the patients’ eyes.

Features from either clinical set A or set B were extracted from the AREDS datasets. In a similar manner to the image features, the clinical features were either concatenated when analyzing multiple visits and the training sets were used to fit the same CoxPH model as described above. No PCA was used for clinical features. Survival analysis only included clinical features or “deep-features.” No datasets contained both.

### 2.7 Hyper-parameter Tuning

Concordance index, ROC AUC at year 5, ROC AUC at year 8, precision-recall AUC at year 5, and precision-recall AUC at year 8 (as described in Evaluation Metrics) on the validation set were calculated for a wide range of learning rates on each combination of the model and dataset using 5-fold cross validation. The best learning rate for each combination was chosen as the learning rate which had the largest product of the mean of these five measures across the 5-fold cross validation in the validation set. All performance metrics shown are evaluated on the independent test sets during cross-validation.

## 3 Results and Discussion

### 3.1 Longitudinal data improves risk prediction with clinical set A

With the limited features available in clinical set A, which are the features used in the 5-step simplifies severity scale, incorporating longitudinal data improves predictive performance (Table 1). Here, we see that the CoxPH with visits at years 0, 2, and 3 performs better than the CoxPH model using only data from year 3. We see a similar result with the MLP, where incorporating longitudinal data improved on the performance in comparison to using the single time point. Interestingly, the concatenated features performed better than the LSTM. As a whole, using longitudinal data with the clinical A dataset seems to be more informative than using just a single time point and the linear model performs equally as well as or better than the deep learning models.

**Table 1:** Results using clinical set A (*: p *<*0.05 from CoxPH (0,2,3))

|  | Concordance Index | ROC AUC@5 | ROC AUC@8 | PR AUC@5 | PR AUC@8 |
| --- | --- | --- | --- | --- | --- |
| CoxPH (3) | 0.869 (0.849, 0.890) | 0.854 (0.820, 0.889) | 0.884 (0.857, 0.911) | 0.075 (0.044, 0.107) | 0.300 (0.247, 0.353) |
| CoxPH (0, 2, 3) | 0.880 (0.864, 0.896) | 0.867 (0.844, 0.889) | 0.893 (0.872, 0.914) | 0.077 (0.049, 0.104) | 0.328 (0.251, 0.405) |
| MLP (3) | 0.862 (0.836, 0.888) | 0.846 (0.795, 0.896) | 0.884 (0.858, 0.910) | 0.082 (0.052, 0.113) | 0.283 (0.241, 0.325) |
| MLP (0, 2, 3) | 0.877 (0.858, 0.896) | 0.868 (0.839, 0.896) | 0.898 (0.881, 0.916) | 0.080 (0.041, 0.119) | 0.328 (0.249, 0.408) |
| LSTM (0, 2, 3) | 0.870 (0.847, 0.892) | 0.860 (0.817, 0.903) | 0.891 (0.874, 0.908) | 0.076 (0.047, 0.105) | 0.319 (0.245, 0.393) |

### 3.2 Deep learning improves risk prediction with clinical set B

Clinical set B proves to be more informative than clinical set A in predicting a patient’s risk of developing late-AMD using all models (Table 2). In the more informative clinical set B, the deep learning models outperform the linear CoxPH model. Dataset B comprises many more features than dataset A, and these features are more “raw,” as in they are evaluated directly by the readers at the grading centers. It is possible that the deep learning models find more indicative interactions between the raw features than the transformation to dataset A does. The binning and dimensionality reduction for dataset A lost a large amount of useful information compared with the 22 features that are provided in dataset B. In this dataset, incorporating multiple timepoints only offers very slight improvements over using the single time point, indicating that the current time point is much more indicative of the patients’ risk of developing late-AMD than the previous time points.

**Table 2:**
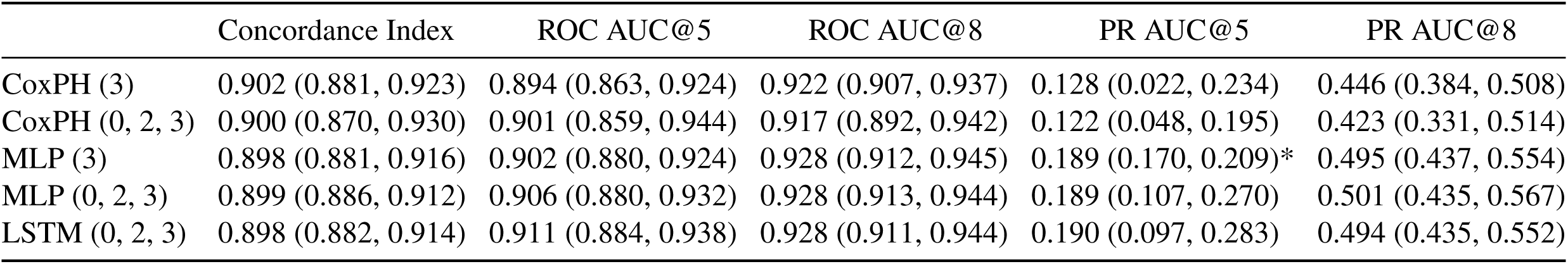
Results using clinical set B (*: p <0.05 from CoxPH (0,2,3))

### 3.3 Pretrained Features are not informative, but incorporating longitudinal data helps

ResNet152 and EfficientNetB3 pretrained features do not perform well (Table 3). The models that extract these features are tuned to classify dogs, birds, boats, and other types of natural image categories. The eye is a very delicate and intricate organ, where small sized abnormalities can cause a large difference. Therefore, the pretrained models are not capable of deciphering the sophisticated features that distinguish a healthy eye from a diseased eye. For both of these feature sets, the deep learning models outperform the CoxPH model. This is especially true in the case of the ResNet152 pretrained features because the PCA was only able to account for 22.4% of the variance in the feature set.

**Table 3:**
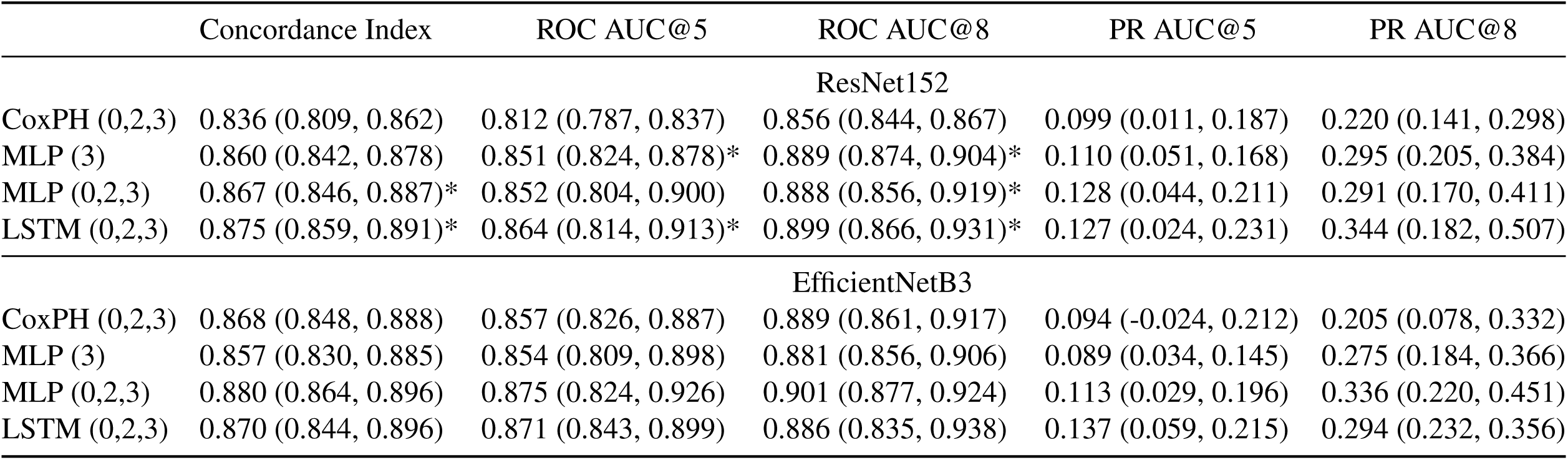
Results using Resenet152 and EfficientNetB3 pretrained on ImageNet (*: p <0.05 from CoxPH (0,2,3))

|  | Concordance Index | ROC AUC@5 | ROC AUC@8 | PR AUC@5 | PR AUC@8 |
| --- | --- | --- | --- | --- | --- |
| ResNet152 |  |  |  |  |  |
| CoxPH (0,2,3) | 0.836 (0.809, 0.862) | 0.812 (0.787, 0.837) | 0.856 (0.844, 0.867) | 0.099 (0.011, 0.187) | 0.220 (0.141, 0.298) |
| MLP (3) | 0.860 (0.842, 0.878) | 0.851 (0.824, 0.878)* | 0.889 (0.874, 0.904)* | 0.110 (0.051, 0.168) | 0.295 (0.205, 0.384) |
| MLP (0,2,3) | 0.867 (0.846, 0.887)* | 0.852 (0.804, 0.900) | 0.888 (0.856, 0.919)* | 0.128 (0.044, 0.211) | 0.291 (0.170, 0.411) |
| LSTM (0,2,3) | 0.875 (0.859, 0.891)* | 0.864 (0.814, 0.913)* | 0.899 (0.866, 0.931)* | 0.127 (0.024, 0.231) | 0.344 (0.182, 0.507) |
| EfficientNetB3 |  |  |  |  |  |
| CoxPH (0,2,3) | 0.868 (0.848, 0.888) | 0.857 (0.826, 0.887) | 0.889 (0.861, 0.917) | 0.094 (-0.024, 0.212) | 0.205 (0.078, 0.332) |
| MLP (3) | 0.857 (0.830, 0.885) | 0.854 (0.809, 0.898) | 0.881 (0.856, 0.906) | 0.089 (0.034, 0.145) | 0.275 (0.184, 0.366) |
| MLP (0,2,3) | 0.880 (0.864, 0.896) | 0.875 (0.824, 0.926) | 0.901 (0.877, 0.924) | 0.113 (0.029, 0.196) | 0.336 (0.220, 0.451) |
| LSTM (0,2,3) | 0.870 (0.844, 0.896) | 0.871 (0.843, 0.899) | 0.886 (0.835, 0.938) | 0.137 (0.059, 0.215) | 0.294 (0.232, 0.356) |

### 3.4 Fine-tuned features are more informative than the features they are trained on

The fine-tuned ResNet152 and EfficientNetB3 outperformed the clinical features and the pretrained features (Table 4). The models were trained on most of the features from dataset A yet outperformed dataset A in a survival setting. This indicates that the deep learning models are able to extract more intricate features than drusen size and pigmentation, which can be more informative in predicting the risk of a patient developing late-AMD than drusen size and pigmentation. Figure 2 displays t-SNE plots of the feature vectors generated from the images used in the survival analysis (years 0, 2, and 3). Individual images are color coded by the amount of time between when that image was taken and when the patient reaches late-AMD. Dark red indicates that the patient is very soon to reach late-AMD, while white dots show that the patient has 8+ years until reaching late-AMD. The gray spots show the overall distribution of image features for both censored and uncensored patients. These t-SNE plots show that the deep feature extractors that are trained on drusen size and presence of pigmentation abnormalities are not only able to predict drusen size and presence of pigmentation abnormalities but are also able to extract features useful in predicting the risk of developing late-AMD without being explicitly trained to do so. These features are not only informative in predicting if a patient will develop late-AMD, but also in predicting if a patient will not develop late-AMD.

**Table 4:** Results using Resenet152 and EfficientNetB3 fine-tuned on AREDS (*: p *<*0.05 from CoxPH (0,2,3))

|  | Concordance Index | ROC AUC@5 | ROC AUC@8 | PR AUC@5 | PR AUC@8 |
| --- | --- | --- | --- | --- | --- |
| ResNet152 |  |  |  |  |  |
| CoxPH (0,2,3) | 0.903 (0.875, 0.930) | 0.901 (0.860, 0.943) | 0.924 (0.897, 0.951) | 0.117 (0.048, 0.185) | 0.327 (0.210, 0.443) |
| MLP (3) | 0.925 (0.898, 0.952) | 0.946 (0.916, 0.976)* | 0.953 (0.927, 0.978) | 0.228 (0.117, 0.340)* | 0.485 (0.340, 0.630)* |
| MLP (0,2,3) | 0.916 (0.887, 0.945) | 0.940 (0.892, 0.988) | 0.941 (0.914, 0.969) | 0.217 (0.079, 0.355) | 0.468 (0.333, 0.602) |
| LSTM (0,2,3) | 0.919 (0.890, 0.949) | 0.946 (0.907, 0.985) | 0.951 (0.925, 0.977) | 0.248 (0.120, 0.375)* | 0.528 (0.444, 0.612)* |
| EfficientNetB3 |  |  |  |  |  |
| CoxPH (0,2,3) | 0.921 (0.899, 0.943) | 0.924 (0.897, 0.952) | 0.942 (0.919, 0.966) | 0.202 (0.142, 0.263) | 0.490 (0.308, 0.673) |
| MLP (3) | 0.929 (0.912, 0.946) | 0.940 (0.906, 0.973) | 0.955 (0.942, 0.968) | 0.244 (0.124, 0.364) | 0.561 (0.436, 0.686) |
| MLP (0,2,3) | 0.925 (0.906, 0.944) | 0.937 (0.906, 0.967) | 0.951 (0.936, 0.966) | 0.216 (0.130, 0.302) | 0.520 (0.384, 0.657) |
| LSTM (0,2,3) | 0.929 (0.911, 0.947) | 0.945 (0.915, 0.975) | 0.954 (0.937, 0.972) | 0.286 (0.157, 0.416) | 0.557 (0.424, 0.691) |

**Table 5:** Results of the multi-task learning classifiers on drusen size (macro-average) and pigment abnormality.

|  |  | ResNet152 | EfficientNetB3 |
| --- | --- | --- | --- |
| Drusen | Overall Accuracy | 0.696 | 0.736 |
|  | Sensitivity | 0.676 | 0.717 |
|  | Specificity | 0.844 | 0.864 |
|  | Precision | 0.681 | 0.735 |
| Pigment Abnormality | Overall Accuracy | 0.876 | 0.853 |
|  | Sensitivity | 0.845 | 0.926 |
|  | Specificity | 0.865 | 0.870 |
|  | Precision | 0.835 | 0.751 |

**Figure 2:**
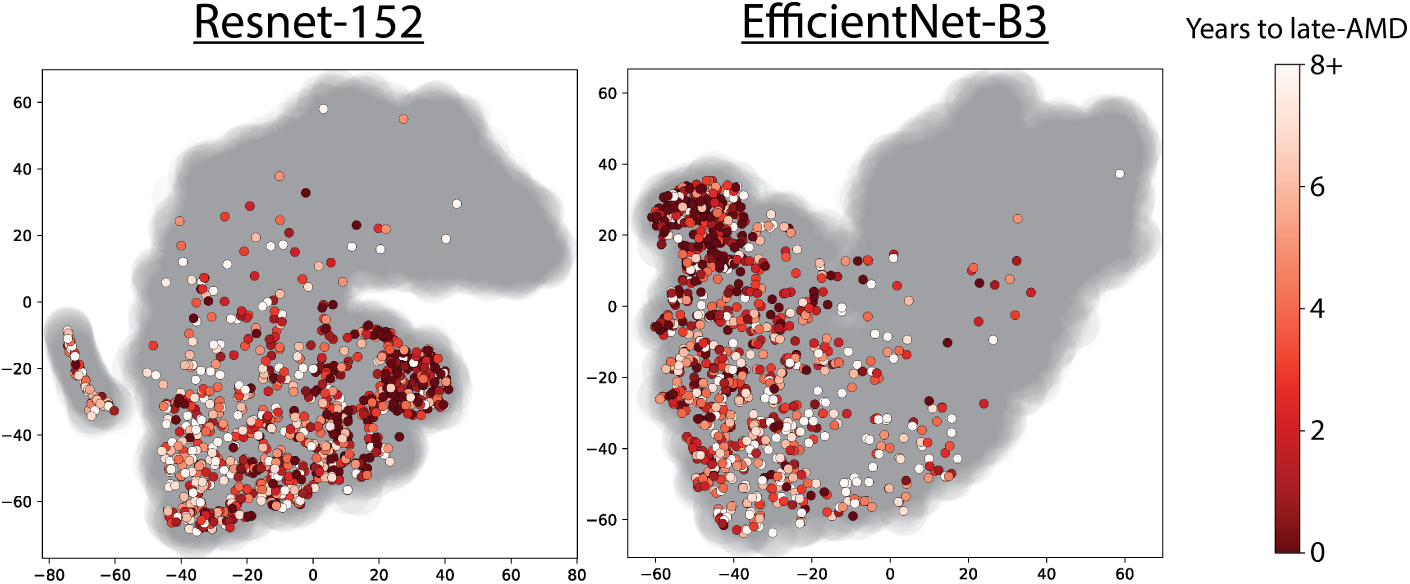
t-SNE plots of the fine-tuned deep features generated from the visits used in the survival analysis (years 0, 2, and 3). Colors indicate time to reach late-AMD. Dark red dots are soon to reach late-AMD. White dots will not reach for many years. Gray spots show the overall distribution of image features censored and uncensored patients.

### 3.5 LSTM improves short term predictions in fine-tuned features

Incorporating longitudinal data from the fine-tuned features does not aid in predicting all time risk of late-AMD. Only the most recent visit is necessary. However, incorporating longitudinal data and modeling the time dependencies of the data using the LSTM model improved on the more immediate prediction, shown by the large increase in PR AUC at year 5 for both the ResNet152 and EfficientNetB3 fine-tuned feature sets.

### 3.6 Multi-task Learning Performance

Both the EfficientNetB3 and ResNet152 were able to accurately classify drusen size and presence of pigmentation abnormalities. The EfficientNetB3 architecture more accurately predicted drusen size than the ResNet152 architecture. Both models had similar accuracies for predicting pigmentation abnormalities. As shown in Figure 3, both of the fine-tuned models are able to accurately cluster the different classifications. Additionally, in both models, we see a gradient from 0 to 2 in predicting drusen size. Class 2 drusen sizes are larger than class 1 and class 0 is the smallest. The models are not explicitly told that this is the case, as each classification is treated as its own binary class. This shows that the fine-tuned CNN models are able to extract valid image features for evaluating fundus photographs.

**Figure 3:**
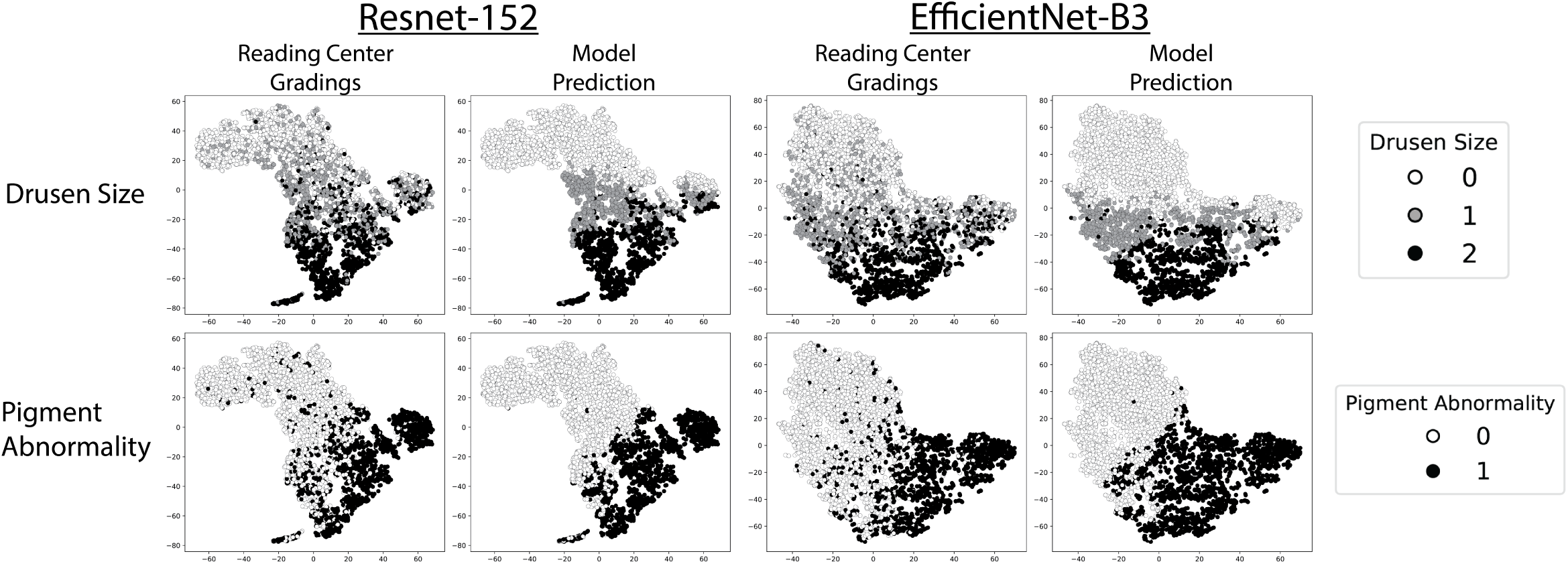
t-SNE plots of the feature vectors generated from the testing set used to evaluate the multi-task learning classifier models. Coloring indicates either the true (Reading Center Gradings) or predicted (Model Prediction) values for classifying drusen size and pigmentation abnormalities.

### 3.7 Discussion

In this study, a multi-task learning framework was used to predict drusen size and pigmentation abnormalities. By visualizing these features using t-SNE and calculating accuracy, sensitivity, and specificity for the model, our results show similar performance to state-of-the-art methods for these classification tasks^8,10^. Interestingly, our features also correlate with the time-to-event prediction, where there was a clear separation for patients that have already reached late stage AMD to those patients who have not. The benefit of using our fine-tuned model, as compared to a pretrained model using Imagenet is shown in the significant increase in model performance based on evaluation metrics in the survival models. In addition, we show that EfficientNetB3, a newer developed model with fewer parameters, as compared to the ResNet152 model, showed higher performance. This may be due to the fact that the EfficientNet models were shown to better capture fine image details within an image^17^. Further, we improved upon the standard CoxPH model that is commonly used in previous literature^11^ using deep learning methods. Last, while incorporating longitudinal data we do see improvements in short-term prediction performance and in clinical feature performance as compared to using a single time point data, but do not see improved long-term performance. It is interesting to note that when year 3 features performed better than years 0-3 features, the data from year 0 showed a decline in model performance (data not shown). This may indicate that more recent time points, for both clinical and image features, are more predictive than earlier time points in predicting risk of developing late-stage AMD.

While the biological changes that occur during AMD are well understood, unfortunately, there is no cure. This model can be used to aid clinicians in predicting how at risk patients are of developing late-AMD. Patients with low risk can have a treatment plan that will decrease patient costs and decrease the burden of patient care on the healthcare system. In contrast, patients with high risk can receive a more aggressive treatment plan, at an earlier point in the disease to prolong vision as long as possible.

This study remains limited. Primarily, of the cohort analyzed in the survival analysis, only 350 patients reached late-AMD and results were not validated on an external dataset. Additionally, measures can be taken to improve the interpretability of the model, such as analyzing saliency maps to see which features are deemed most indicative in the multi-task learning models. To expand the model, the multi-task learning classifier could be extended by increasing the number of tasks learned. Adding in confidence scores would allow this model to aid grading centers by both reducing the time needed to analyze simple cases and assist in edge-case classification. Finally, although we explored a wide range of hyper-parameters and architecture makeups, we plan to conduct a more exhaustive analysis of MLP architectures and other hyper-parameter tunings.

## 4 Conclusion

This study shows that multi-task learning can be used to extract image features that are highly predictive of developing late-AMD. These extracted features are more predictive than the expert grader acquired feature, which are labor intensive and expensive to generate. This model can be used to aid clinicians in the stratification of patients with early-stage AMD, based on the risk of progressing to late-stage AMD. This would ease the exhaustive burden on the experts in the grading centers and greatly reduce cost. Additional future directions include integrating clinical features, such as smoking and age, and image features into the same deep learning model to try to improve model performance.

This model architecture is applicable to many other eye related diseases, including longitudinal prognosis of glaucoma. Additionally, the model could be extended far beyond fundus photographs to aid in longitudinal evaluation of non-eye related diseases such as cancers^27^, COVID-19^28^, and other diseases and illnesses.

## Data Availability

The data is available at https://www.ncbi.nlm.nih.gov/projects/gap/cgi-bin/study.cgi?study_id=phs000001.v3.p1

https://github.com/bionlplab/AMD_prognosis_amia2021

## Data Availability

https://github.com/bionlplab/AMD_prognosis_amia2021

## Data Availability

https://github.com/bionlplab/AMD_prognosis_amia2021

## Acknowledgements

The work was supported by the intramural program funds and contracts from the National Center for Biotechnology Information/National Library of Medicine/National Institutes of Health, the National Eye Institute/National Institutes of Health, Department of Health and Human Services (Contract HHS-N-260-2005-00007-C; ADB contract NO1-EY-5-0007; Grant No 4R00LM013001; NSF 1750326; NIH NIMH R01MH124740; NIH NIA RF1AG072449).

## Footnotes

c https://www.ncbi.nlm.nih.gov/projects/gap/cgi-bin/study.cgi?study_id=phs000001.v3.p1

d https://imgaug.readthedocs.io/en/latest/index.html

e https://github.com/rwightman/gen-efficientnet-pytorch

f https://github.com/CamDavidsonPilon/lifelines

g https://cran.r-project.org/web/packages/timeROC/timeROC.pdf

h https://scikit-learn.org

i https://matplotlib.org/

